# Evolving SARS-CoV-2 virulence among hospital and university affiliates in Spain and Greater Boston

**DOI:** 10.1101/2022.11.28.22282412

**Authors:** Fares Amer, Fan-Yun Lan, Mario Gil-Conesa, Amalia Sidossis, Daniel Bruque, Eirini Iliaki, Jane Buley, Neetha Nathan, Lou Ann Bruno-Murtha, Silvia Carlos, Stefanos N. Kales, Alejandro Fernandez-Montero

## Abstract

**Background:** The COVID-19 pandemic caused by the SARS-CoV-2 virus greatly affected healthcare workers and healthcare systems. It also challenged schools and universities worldwide negatively affecting in-person education. We conducted this study is to assess the evolution of SARs-CoV-2 virulence over the course of the pandemic.

**Methods:** A combined cohort of affiliates from the University of Navarra, two hospitals in Spain, and one healthcare system in the Greater Boston area was followed prospectively from March 8^th^, 2020, to January 31^st^, 2022 for diagnosis with COVID-19 by PCR testing and related sequelae. Follow-up time was divided into four periods according to distinct waves of infection during the pandemic. Severity of COVID-19 was measured by case-hospitalization rate. Descriptive statistics and multivariable-adjusted statistics using the Poisson mixed-effects regression model were applied.

**Results:** For the last two periods of the study (January 1^st^ to December 15^th^, 2021 and December 16^th^, 2021 to January 31^st^, 2022) and relative to the first period (March 8^th^ to May 31^st^, 2020), the incidence rate ratios (IRRs) of hospitalization were 0.08 (95% CI, 0.03-0.17) and 0.03 (95% CI, 0.01-0.15), respectively.

**Interpretation:** The virulence of COVID-19 and immunity of our populations evolved over time, resulting in a decrease in case severity. We found the case-hospitalization rate decreased more than 90% in our cohort despite an increase in incidence.

## Introduction

Year to date (July 2022), the World Health Organization has confirmed more than 515 million cases of COVID-19 over the world, more than 6.25 million deaths and more than 11,500 million vaccine doses administered. The first wave of the pandemic in the winter/spring of 2020 brought an oversaturation of hospitals and ICUs, extreme fatigue to healthcare workers (HCWs) and a lack of Personal Protective Equipment (PPE) due to the huge increase of the demand. This situation left HCWs more susceptible to SARS-CoV-2 (the virus causing COVID-19) and led to more than 115,000 deaths between March 2020 and May 2021^1^.

Since the first outbreak of SARS-CoV-2 in December 2019 with the Alpha variant, the virus has mutated into more than 10 variants, reaching the current Omicron variant predominance since December 2021. These variants differ in their speed of emergence, transmissibility and virulence^2^, which along with population immunity they determine the impact on the population in terms of COVID-19 severity. In fact, previous literature has shown decreasing COVID-19 complications throughout the first few months after the initial outbreak^3^. On the other hand, the pharmaceutical industry urgently began to develop treatments^4^ and vaccines against SARS-CoV-2 using different mechanisms^5^ while at the hospitals some existing medication was used, such as azithromycin, hydroxychloroquine or dexamethasone^6^, trying to mitigate the disease severity. COVID-19 vaccines were designed to protect against SARS-CoV-2, while evidence has shown the effectiveness against different variants varies. In general, the primary vaccination series and natural immunity have reduced the severity and death rates of the disease^2^. The evolution of the virus, an accumulation of knowledge, population immunity and the development of treatments have altogether led to a better prognosis of COVID-19 over time.

While school and university closures were intended to mitigate the transmission of SARS-CoV-2^7^, many negative social and educational impacts have been reported. These include the disruption of learning continuity, a lack of social interaction with teachers and peers, aggravation of disparities, worse academic performance^8^ and burnout^9^. Therefore, evidence supports keeping schools and universities open.

The aim of this study was to evaluate the evolution in virulence of SARS-CoV-2 infection over the course the pandemic as measured by case-hospitalization rates, among HCW and a Spanish university that remained open, with face-to-face teaching.

## Materials and methods

Between March 8^th^, 2020 and January 31^st^, 2022, a prospective cohort study was conducted on members of a study base who were diagnosed with SARS-CoV-2 infection by PCR testing. Cases were drawn from a total of 22,405 individuals affiliated with the University of Navarra in Spain (Students and employees) and healthcare professionals from two different healthcare systems in three cities: Clínica Universidad de Navarra (CUN) in Pamplona and Madrid and Cambridge Health Alliance (CHA) in the Greater Boston area.

### Variables

We categorized the main exposure variable of interest, timing throughout the pandemic, into four periods. The first period corresponds to COVID-19 pandemic’s first wave from March 8^th^ to May 30^th^, 2020; the second period corresponds to the time prior to primary vaccination from June 1^st^ to December 31^st^, 2020; the third period covers primary vaccination campaigns and delta variant predominance from January 1^st^ to December 15^th^, 2021; and the fourth period corresponds to the delivery of booster doses and the emergence of Omicron variant from December 16^th^, 2021 to January 31^st^, 2022. The main outcome measure of COVID-19 severity was case-hospitalization rate. Other outcomes included the rates of intubations and deaths. In addition, we collected the cases’ sociodemographic variables such as age, sex and workplace, as well as reinfection events. On the other hand, we collected SARS-CoV-2 RNA information in Boston-area wastewater provided by Biobot Analytics (https://www.mwra.com/biobot/biobotdata.htm), and information on COVID-19 deaths in the same area and in the same period (https://www.mass.gov/info-details/covid-19-response-reporting).

### Statistical analysis

For descriptive analyses of continuous variables, means, standard deviations and 95% confidence intervals were used, and t-tests were performed. The Chi-square test or Fisher’s exact test, as appropriate, was used to compare the categorical variables across the four periods of the study. The mixed-effects Poisson regression model adjusting for sex, age, work-center, healthcare worker status and reinfection was conducted to evaluate the hospitalization risk for each period of time. COVID-19 incidence was calculated as cumulative incidence in 14 days (CI14 = number of new COVID-19 cases during a 14-day period prior to and including the given date). We accounted for correlated data since reinfected cases were included in the analysis and contributed to more than one data entries.

Finally, crude relative risks of death from COVID-19 and SARS-CoV-2 RNA data in Boston-area wastewater were calculated as a sensitivity analysis, using the first wave as the reference category. A two-tailed p values < 0.05 were considered to be statistically significant. The STATA 15 and R software 3.6.3 were used for statistical analysis.

The study was conducted following the STROBE guidelines for cohort studies and in compliance with the study protocol, the current version of the Declaration of Helsinki, and the local, legal and regulatory requirements (Approved by the University of Navarra Ethics committee: 2020.190 on October 30th, 2020 and the Cambridge Health Alliance Institutional Review Board (4/29/202-003)).

## Results

The total incidence of COVID-19 throughout the four periods was 5,710 PCR-confirmed cases, where 156 cases were reinfections. The mean age of the study population was 31.7 years old (95% CI: 18.5-44.9), with mostly women, 67%. The cases’ characteristics and outcomes are described in Table 1, stratified by the four time periods.

**Table 1.** Baseline characteristics of COVID-19 positive cases across the time periods 8^th^ March 2020 – 31^st^ January 2022.

| Time periods |  | 8<sup>th</sup> March 2020<br>to 31<sup>st</sup> May 2020 | 1<sup>st</sup> June 2020 to<br>31<sup>st</sup> December<br>2020 | 1<sup>st</sup> January 2021<br>to 15<sup>th</sup> December<br>2021 | 16<sup>th</sup> December<br>2021 to 31<sup>st</sup><br>January 2022 | p |
| --- | --- | --- | --- | --- | --- | --- |
| Positive cases, n |  | 309 | 918 | 1438 | 3045 |  |
| Age, years (mean±SD) |  | 42.7 ±12.3 | 32±13.7 | 29.9±12.2 | 31.5 ±13.1 | <0.001 |
| Sex, male (%) |  | 68 (22.0) | 332 (36.2) | 498 (34.6) | 951 (31.2) | <0.001 |
| Workplace (%) | UNAV | 0 (0) | 556 (60.6) | 1003 (69.7) | 1870 (61.4) | <0.001 |
|  | CUN Pamplona | 97 (31.4) | 37 (4.0) | 99 (6.9) | 367 (12.1) |  |
|  | CUN Madrid | 54 (17.5) | 46 (5.0) | 59 (4.1) | 29 (1.0) |  |
|  | CHA | 158 (51.1) | 279 (30.4) | 277 (19.3) | 779 (25.6) |  |
| Healthcare workers (%) |  | 309 (100) | 362 (39.4) | 435 (30.3) | 1175 (38.6) | <0.001 |
| Reinfection (%) |  | 0 (0) | 2 (0.2) | 12 (0.8) | 142 (4.7) | <0.001<sup>a</sup> |
| Hospitalization (%) |  | 18 (5.8) | 24 (2.6) | 14 (1) | 2 (0.1) | <0.001<sup>a</sup> |
| Intubation (%) |  | 1 (0.3) | 1 (0.1) | 0 (0) | 0 (0) | 0.020<sup>a</sup> |
| Death (%) |  | 1 (0.3) | 1 (0.1) | 0 (0) | 0 (0) | 0.020<sup>a</sup> |

Mean ±SD for age. Count (%) for other variables.
UNAV; University of Navarra
CUN Pamplona; University of Navarra Clinic, located in Pamplona
CUN Madrid; University of Navarra Clinic, located in Madrid
CHA; Cambridge Health Alliance
<sup>a</sup> Derived from Fisher's exact test

In the first period, all the positive cases were HCWs because all university students and employees stayed at home due to the mandatory lockdown implemented in Spain starting on March 14^th^. In the following periods, CHA center HCWs represented between 20% and 30% of positive cases, and all infected HCWs represented between 30% and 40% of the total number of cases. The greatest number of positive COVID-19 cases, more than 60%, corresponded to the University of Navarra students and employees.

The observed crude Incidence Rate Ratio (IRR) of hospitalization among COVID-19 cases, or case-hospitalization rate ratio, in the second, third and fourth periods compared with the first period was 0.2 (95% CI: 0.11-0.37), 0.03 (95% CI: 0.01-0.05) and 0.01 (95% CI: 0.003-0.06), respectively (Table 2). After adjusting for several factors (i.e., sex, age, work center, healthcare worker status and reinfection), the decrement of hospitalization risk for the second period became non-significant, while the third and fourth period presented a significant decrease in hospitalizations, with an IRR of 0.08 (95% CI: 0.03-0.17) and 0.03 (95% CI: 0.01-0.15), respectively.

**Table 2.** Hospitalization risk according to time periods 8th March 2020 – 31st January 2022.

| Hospitalization | 8<sup>th</sup> March 2020<br>to 31<sup>st</sup> May<br>2020 | 1<sup>st</sup> June 2020 to<br>31<sup>st</sup> December<br>2020 | 1<sup>st</sup> January 2021<br>to 15<sup>th</sup> December<br>2021 | 16<sup>th</sup> December<br>2021 to 31<sup>st</sup><br>January 2022 |
| --- | --- | --- | --- | --- |
| Events/ Person-days at risk<sup>a</sup> | 18/17309 | 24/116224 | 14/498729 | 2/137287 |
| Crude IRR<sup>b</sup> (95% CI) | 1 (ref.) | 0.20 (0.11-0.37) | 0.03 (0.01-0.05) | 0.01 (0.003-0.06) |
| Sex and age adjusted IRR<sup>b</sup> (95% CI) | 1 (ref.) | 0.42 (0.22-0.82) | 0.07 (0.03-0.14) | 0.03 (0.01-0.13) |
| Multivariable-adjusted IRR<sup>bc</sup> (95% CI) | 1 (ref.) | 0.57 (0.27-1.21) | 0.08 (0.03-0.17) | 0.03 (0.01-0.15) |

<sup>a</sup> Each case was followed from the date of diagnosis until the end of the period or death, whichever came first.
<sup>b</sup> IRR: Incidence rate ratio; derived from the mixed-effects Poisson regression model accounting for correlated data and person-days.
<sup>c</sup> Adjusted for sex, age, work-center, healthcare worker status and reinfection.

## Discussion

In this prospective cohort study, we followed HCWs of two different healthcare systems in the USA and Spain as well as affiliates with the University of Navarra in Spain between March 8^th^ 2020 and January 31^st^ 2022. We found a 97% reduction in hospitalization risks comparing the last period to initial outbreak, while the number of cases increased almost 10 times throughout the study period. Our novel findings provide real-world evidence of evolving SARS-CoV-2 virulence, which has decreased dramatically over time as the population gained widespread immunity and the virus became an endemic infection.

In the first period starting from March 14^th^ 2020, despite mandatory lockdowns we observed the highest rate of case-hospitalization rate, 5.8%. Likely explanations include: a largely immunologically naïve population, a more virulent strain of virus, and less initial knowledge regarding transmission, protective measures and effective treatments.

Since the onset of the second period, research and practice identified various SARS-CoV-2 treatments such as corticosteroids, hydroxychloroquine, remdesivir, monoclonal antibodies, vaccines and lopinavir/ritonavir to mitigate severe COVID-19 complications^10,11^. During this period, in October 2020, Delta variant was first identified and subsequently became globally dominant in June 2021. Compared to the original strain, the Delta variant was more contagious with a 97% higher transmission rate^12^. Accordingly, we found an increase of COVID-19 cases in our population, while observed a less severe health impact (almost 50% less of hospitalizations in comparison with the first period) and a softer criteria for hospital admissions.

The first COVID-19 vaccine that received the emergency use authorization (EUA) was Pfizer’s, in November 2020 in both the USA and Europe, and the vaccination campaigns started soon in December 2020. Therefore, the populational immunity had been rapidly established during the third period, especially among HCWs who were prioritized in the campaigns. In fact, most people in the study got their first dose of vaccine between December 2020 and June 2021. Even though the Delta variant became predominant in the summer of 2021, with its greater transmissibility and ability to escape immunity, we still observed a continued decreasing trend in complications.

The fourth period was characterized by the spread of the Omicron variant, which is from 3 to 5 times less virulent than the Delta variant, but more contagious13. Nonetheless, we continued to observe decreased hospitalizations. Overall, throughout the study period, the incidence of COVID-19 and its CI14 increased over time, especially in certain periods such as the first wave in March 2020, the beginning of the academic year in 2020, Christmas Eve in 2020, Easter in 2021 and Christmas Eve in 2021. However, we observed that hospitalizations decreased, despite an increasing disease incidence and its CI14 (Figure 1A and 1B). To provide robustness to these claims, we calculated the relative risk of deaths from COVID-19 in relation to the amount of SARS-CoV-2 RNA measured in wastewater and observed that the risks of deaths decreased from the first pandemic period to a RR= 0.009 CI95% (0.009-0.010) in the fourth period. (Figure 2A and 2B)

**Figure 1.**
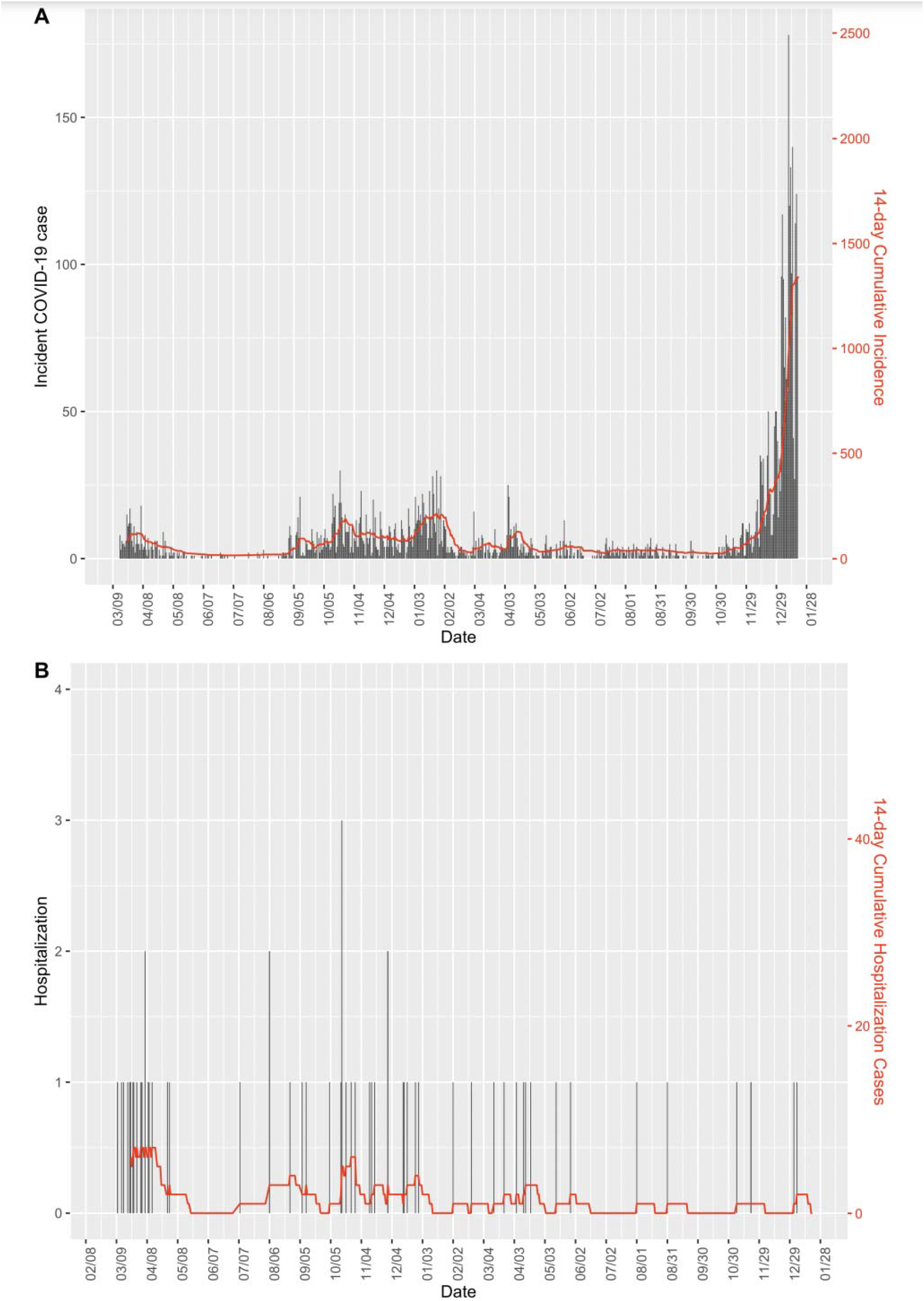
Cumulative incidence in 14 days (CI14) between March 2020 and January 2022. A.) COVID-19 cases. B.) Hospitalization COVID-19 cases.

**Figure 2.**
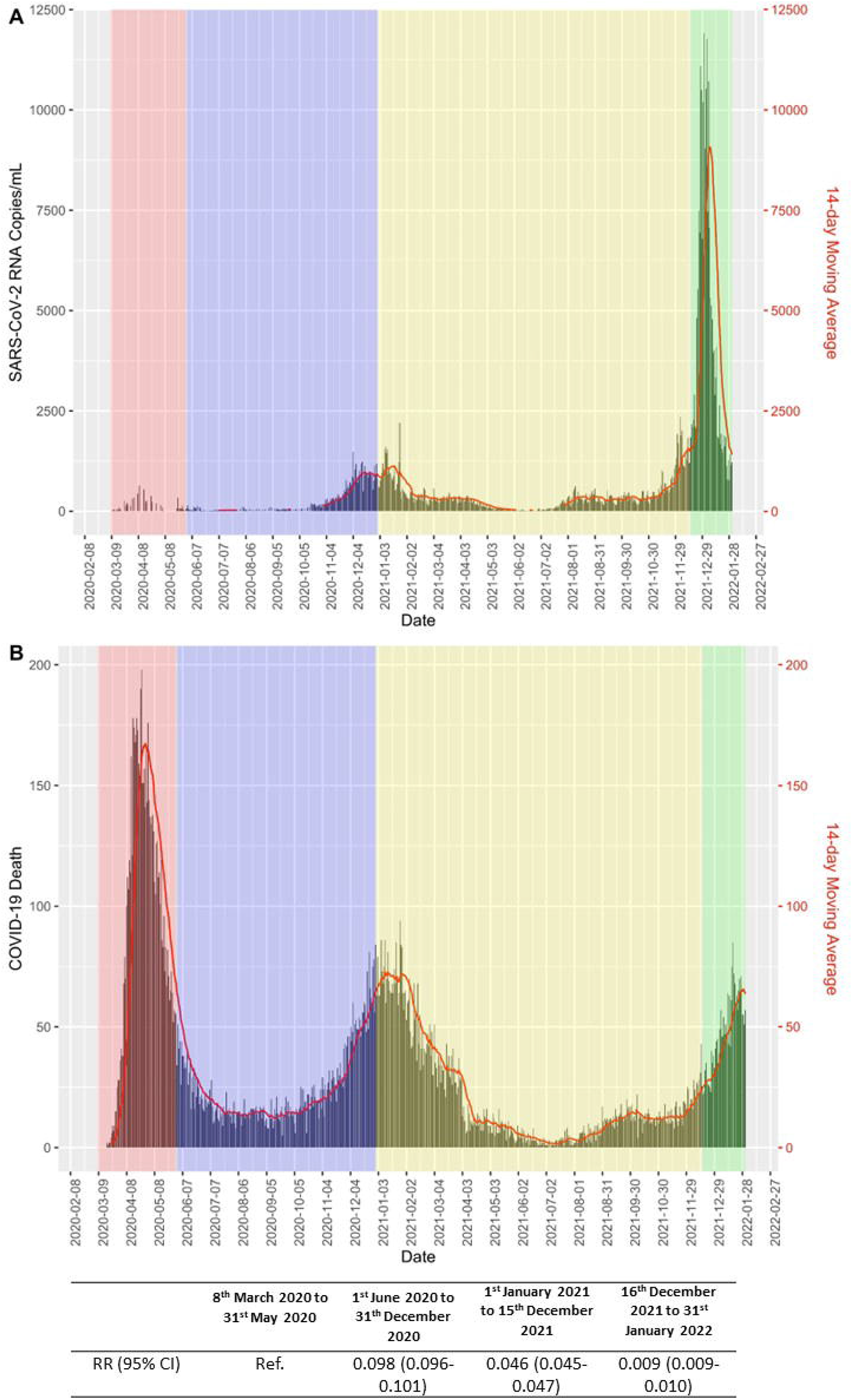
**Cumulative incidence in 14 days (CI14) between March 2020 and January 2022. A.) SARS-CoV-2 RNA Copies/mL** in Boston-area wastewater. **B.) Death of COVID-19 cases** in Boston-area. **Relative Risk and 95%CI between the two measures.**

Our study has some limitations. First, our cohort was subject to a healthy worker/student effect relative to the general population, and thus, we had few cases of hospitalization, death and ICU admission. The small number of the outcomes limited our statistical power to analyze the risk of death and ICU admission. In addition, we did not collect information on symptoms at the time of hospital admission, and it is likely that at the later period, when more contagious variants emerged, COVID-19 hospitalization could be misclassified due to incidental findings from regular admission screening programs. The differential information bias could make our findings an underestimation. Finally, we cannot tell from the results whether the decreased disease severity was due to viral evolution, vaccination rate, healthcare improvement, or other factors. However, the observation of more mild cases and less severe complications over time is a result of all these mechanisms combined. Nonetheless, the study does have several strengths. First, all COVID-19 cases were diagnosed by PCR and were not self-referred, freeing the study population from misclassification. In addition, there is a large sample of 5,710 incident cases across USA and Spain, which allowed us to analyze the risk of hospitalization in the different periods and enhance generalizability. Finally, all cases’ information was provided directly by the human resources department or extracted from the official diagnostic database of each institution, minimizing information bias. In conclusion, our present study on an international cohort of HCWs and university affiliates demonstrates that SARS-CoV-2 virulence measured in number of hospitalizations and deaths has reduced by 98% and 100%, respectively, during the later periods of the pandemic.

## Data Availability

All data produced in the present study are available upon reasonable request to the authors

## Data availability statement

The data set is available to any interested reader. Please contact the corresponding author at

